# The trends in maternal age distribution and the estimated live birth and population prevalence of Down’s syndrome in China: 1985-2012

**DOI:** 10.1101/2021.04.05.21254264

**Authors:** Jing Kang, Jianhua Wu

## Abstract

This study aims to investigate the trends in maternal age distribution and estimate the live birth and population prevalence of Down’s syndrome in China.

Using population survey data, we demonstrated the change of maternal age over the past three decades and its effect on the live birth prevalence of Down’s syndrome. We also integrated the live birth prevalence and the survival rate to estimate the population prevalence of Down’s syndrome. Chi-square test was used to compare the maternal age distributions across survey years.

The results show that the maternal age has been rising over the past 30 years in China at national level. The proportion of mothers aged 35 and over increased from 3.62% in 1985 to 14.40% in 2010. The estimated live birth prevalence of Down’s syndrome has mirrored the same increase of the maternal age from 1.07 per 1000 live births in 1985 to 2.36 per 1000 live births in 2010. At City level, the change of maternal age and live birth prevalence of Down’s syndrome were more significant than at Town and County levels. The proportion of mothers aged 35 and over increased from 2.17% in 1995 to about 16% in 2010 at City level, while it increased from 2.03% to 13.65% at County level. The total estimated number of people with Down’s syndrome who were still alive in 2012 and aged below 27 was 611,053, and the estimated population prevalence is 0.45 per 1000.

To conclude, this study provides a clear message about the rising trend in maternal age in both urban and rural areas in China since 1985 and its effect on the estimated birth prevalence with Down’s syndrome. The regional differences should be taken into account for planning population policy and allocating medical resources to reduce the burden of family with Down’s syndrome.

## INTRODUCTION

Down’s syndrome is the most common genetic disorders in humans, and its prevalence has been extensively studied and reported. Data for the live birth children with Down’s syndrome have been recorded by national registries in the developed countries such as the UK (1, 2) and US (3) since the late 80s, but such national data is yet available in the fast developing countries such as China (4). The live birth prevalence of babies with Down’s syndrome has been reported in several regions in China, but the vast regional and economical differences across China and the lack of national birth data makes it difficult to estimate the population birth prevalence of Down’s syndrome.

The risk of live birth with Down’s syndrome in the absence of antenatal screening is closely associated with maternal age, and the association formula developed within western population has found to be not significantly different from the Chinese population(4). Therefore the maternal age distribution could be used to predict the birth prevalence of Down’s syndrome in the absence of antenatal screening. Unlike the stabilized age distribution in western developed countries, the age distribution in China has kept changing over the past few decades. The one-child policy implemented about 30 years ago, the fast economic growth, changing lifestyle, and recently opened second-child policy all have profound impact on the maternal age and have shaped the maternal age distribution over the years. Moreover, in recent years couples tend to have children at later age due to financial and career pressures.

All these factors could contribute to the livebirth prevalence of Down’s syndrome in the absence of antenatal screening.

This paper aims to investigate the trends in maternal age distribution in China from 1985 to 2012, and estimate the live birth prevalence of Down’s syndrome in the absence of antenatal screening. The study also aims to estimate the population prevalence of Down’s syndrome in 2012 by integrating the livebirth prevalence and the survival rate of Down’s syndrome.

## MATERIALS AND METHODS

### Maternal age

The maternal data was obtained from China Population Statistical Yearbook (5) published by National Bureau of Statistics since 1985 onwards. China Population Statistical Yearbook is an annual statistical publication containing figures of the family planning statistics on the maternal age and the live births at the national level and local levels of province and autonomous regions of the past year. The Yearbook produced statistics from population survey data collected through multistage stratified clustering survey sampling techniques with selection probability proportionate to population size. The data in 1990, 2000 and 2010 were produced through population census, while the 1985, 1995 and 2005 data were sampled at the proportion of 1/100. The data in other years were sampled at the rate of 1/1000. The data presented in the Yearbook tables were weighted and meant to represent the population of China of that year proportionately.

The maternal age and the number of live births were extracted from the Yearbook between 1985 and 2012 at national level as well as categorized according to administrative divisions (City, Town and County). The number of live births was grouped by mother’s age at birth (years). Chi-square test was used to compare the maternal age distribution across different administrative divisions (Country, City, Town, and County) and years (1995, 2000, 2005, and 2010).

### Live birth and population prevalence of Down’s syndrome

The risk of Down’s syndrome is mainly affected by the maternal age and the association can be approximately estimated with the following equation (6, 7)

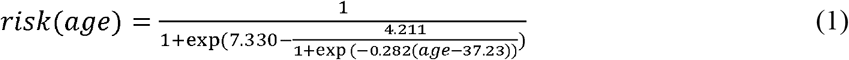

The number of live births with Down’s syndrome for a given maternal age was estimated by integrating the risk of Down’s syndrome in equation (1) with the number of live births for that given maternal age. The live birth prevalence of Down’s syndrome for a given year was estimated by dividing the total number of live births with Down’s syndrome by the total number of live births of the year.

This paper used the survival rate for people with Down’s syndrome at different age fitted by Wu and Morris in 2013 (1) for western population. For any live births with Down’s syndrome before 2012, the probability that infant survived to 2012 could be calculated. Therefore the total number of people survived to 2012 for any given year could be estimated by integrating the number of live births and the survival probability for that given year. The number of people with Down’s syndrome in 2012 was the sum of people of all ages with Down’s syndrome who were still alive.

## RESULT

### Maternal age distribution

The age-specific risk for Down’s syndrome is plotted against maternal age (15-45) in figure 1. The risk for Down’s syndrome is fairly small (less than 3 per 1000) for maternal age below 35, however the risk increases significantly to 28 per 1000 at age of 45 years. The risk of Down’s syndrome for age over 45 years plateaued (6), thus we replaced the risk for age over 45 years with the risk at age 45.

**Figure 1:**
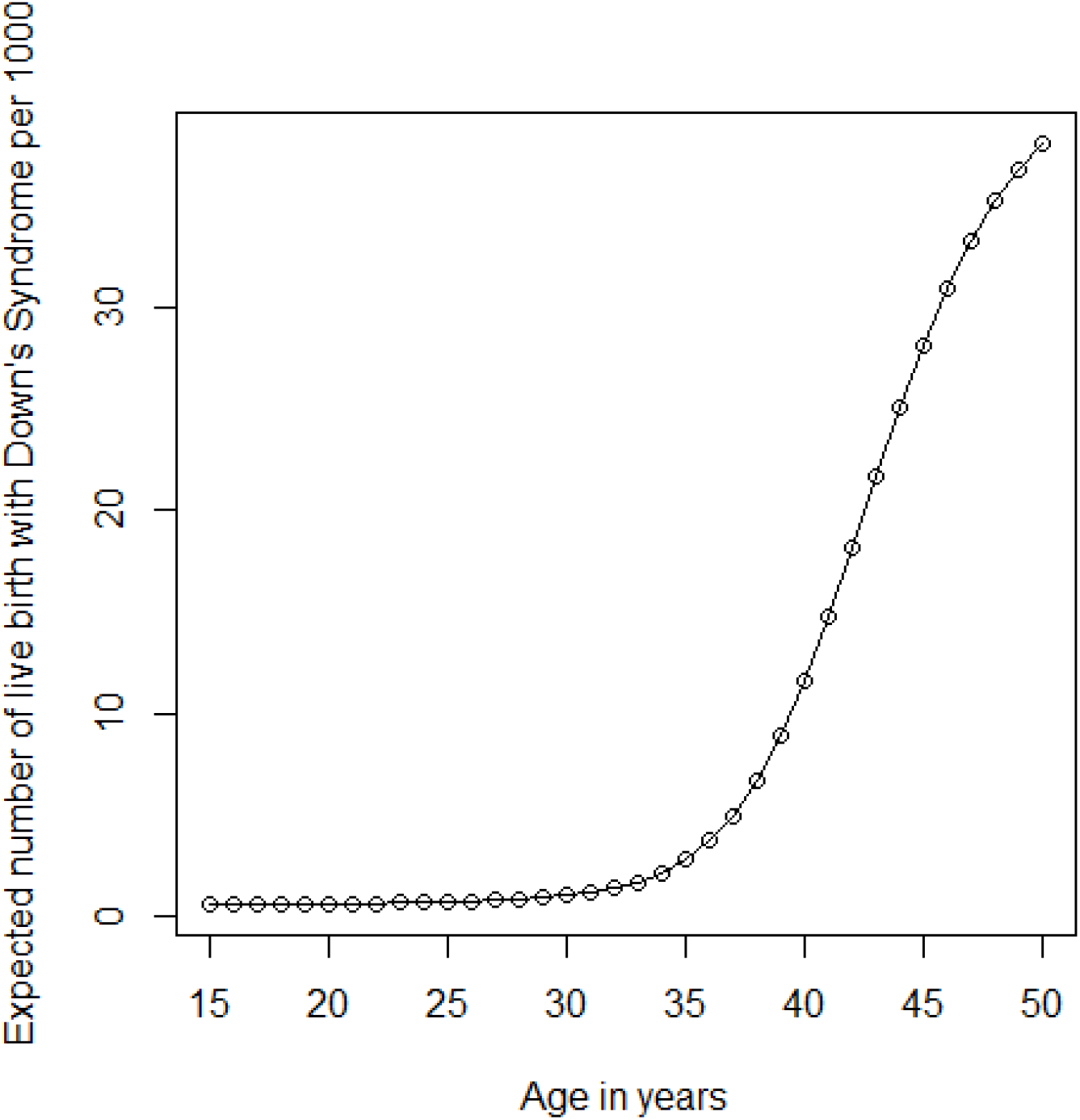
Expected risk of babies with Down’s syndrome per 1000 live births against maternal age (15 to 45).

The maternal age distribution has changed significantly from 1985 to 2010 nationally (Figure 2a, supplement table 1 and Supplement figure 1). The national maternal age distribution was significantly different between 1985 and 2000 (chi-square test, p-value < 0.001), and between 2000 and 2010 (chi-square test, p-value < 0.001). The proportion of mothers aged under 25 kept decreasing continually from 58% in 1985 to 35% in 2010, while the proportion of mothers aged 35 and over stayed around 4% between 1985 and 2000, and increased to about 14.4% in 2010. Such changes of pattern in maternal age were also reflected at different administrative divisions (figure 2b-d, supplement table 1 and Supplement figure 1).

**Figure 2.**
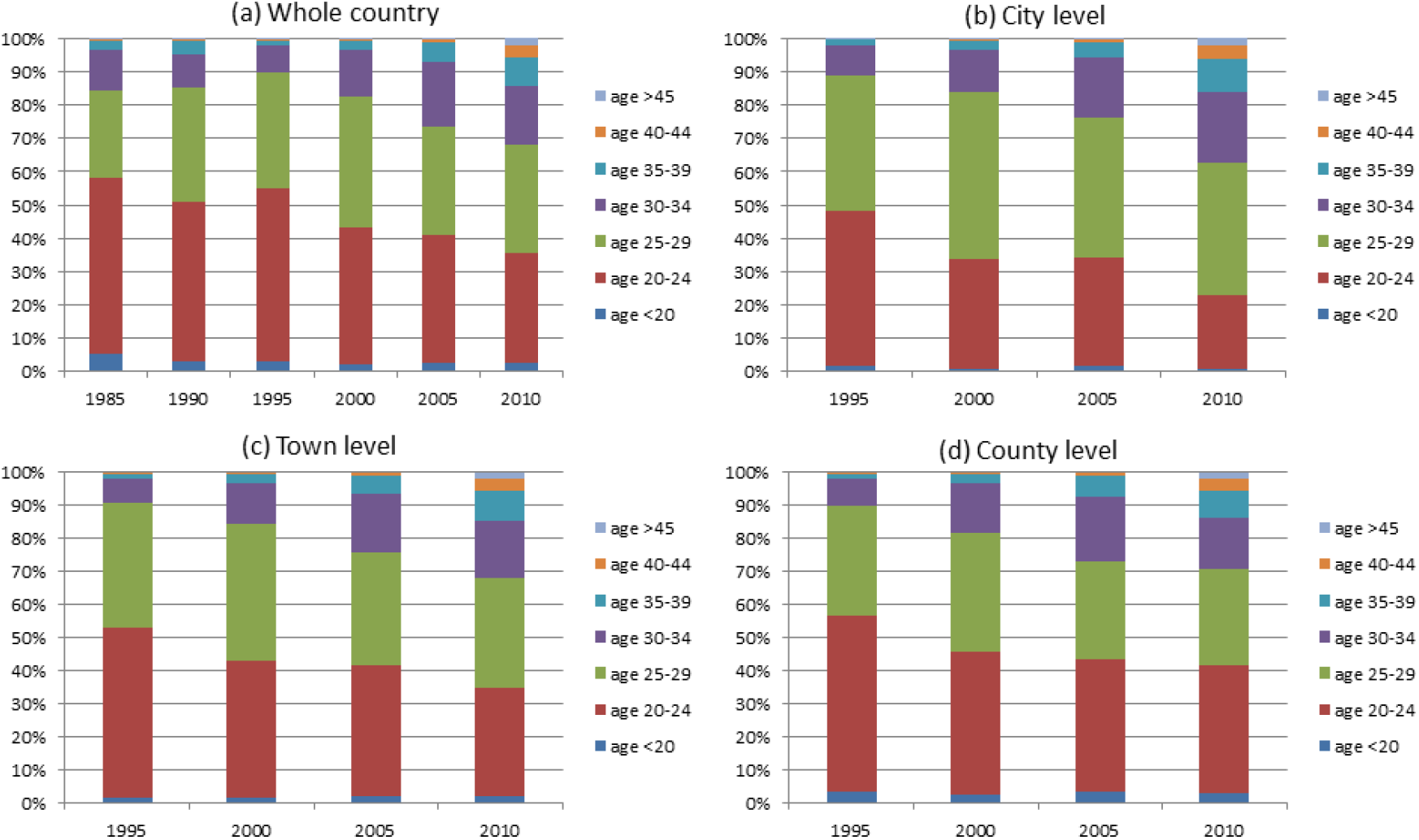
Maternal age distribution in China between 1985 and 2010 at national scale (a) and in different administrative regions (city, town and county, panels b-d).

We compared the maternal age distribution across four survey years (1995, 2000, 2005 and 2010) as well as administrative divisions (City, Town and County). The maternal age distribution in regions of city, town and county are significantly different at any particular year (all p-values < 0.001). For example, when comparing between city and county in 2010, the proportion of mothers aged under 25 was 42% for county and 22% for city. But when comparing the age distribution for city between two survey years 1995 and 2010, the proportion of mothers aged under 25 was 48% in 1995 and 22% in 2010.

The proportions of mothers aged 35 and over at country and regional levels are displayed in figure 3 for year 1985-2012. The proportion of mothers aged 35 and over stayed the same at about 5% from 1985 to 2000, then increased significantly to reach a peak of 16.7% in 2008 at both national and regional levels, and started to drop slightly.

**Figure 3.**
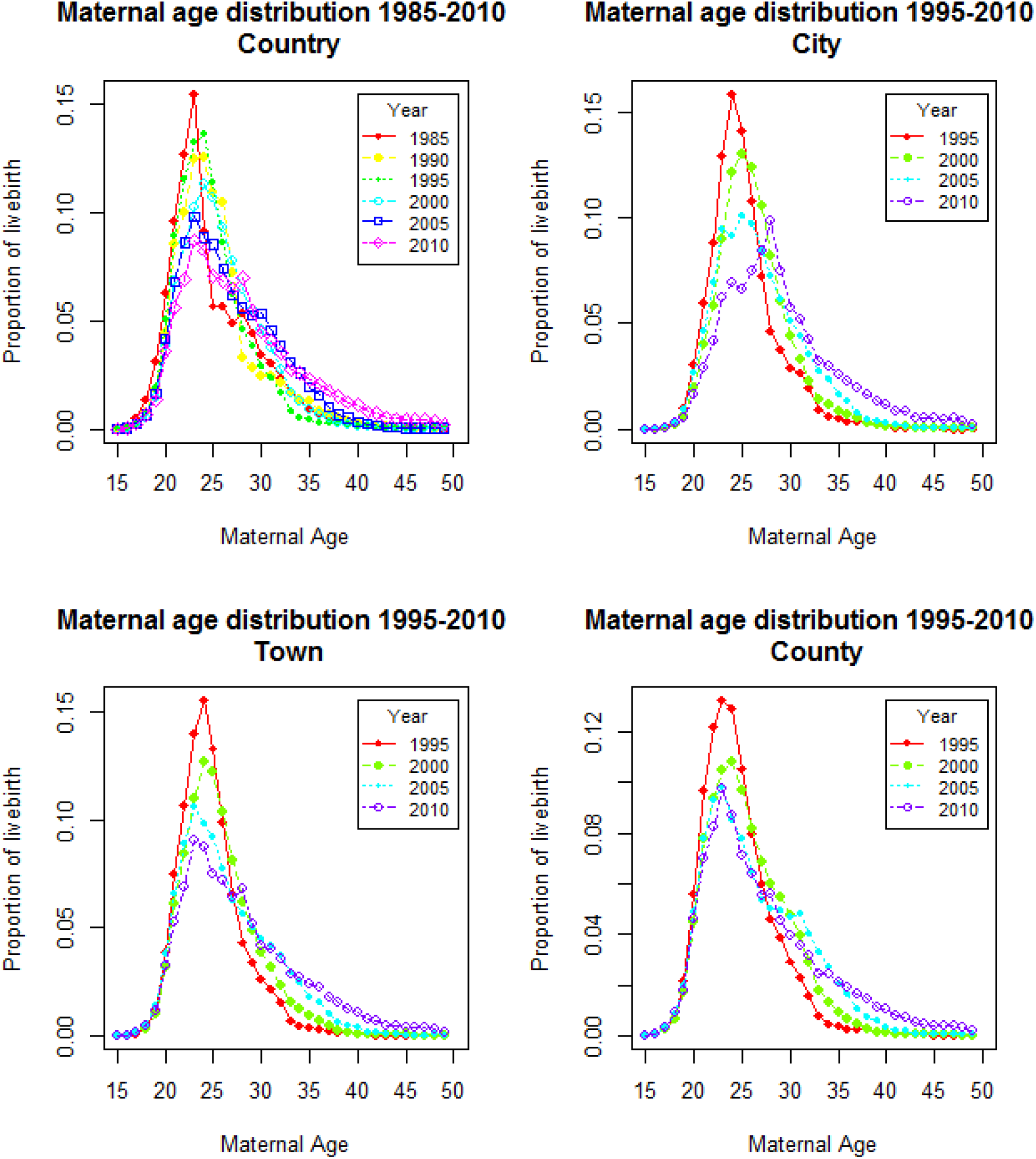
Proportion of mothers aged 35 and over and the live birth prevalence of Down’s syndrome from 1985 to 2012 for country (panel a) and administrative regions (city, town and county, panels b-d).

### Live birth prevalence of Down’s syndrome

The total number of live births with Down’s syndrome in China, in the absence of prenatal diagnosis and subsequent terminations, was estimated by integrating the DS risk by age in equation (1) with the number of live births in that age and summing across all ages, as no program in China has collected actual number of babies born with Down’s syndrome, comparing with the UK National Down’s Syndrome Cytogenetic Register (7) started in 1989. The estimated live birth prevalence of Down’s syndrome mirrored the trend of the proportion of mothers aged 35 and over (Figure 3) with a peak of 2.71 per 1000 livebirths nationally at 2008 (Figure 3a), and as high as 3.72 per 1000 live births in City in the same year. The prevalence of Down’s syndrome in County was smaller than that in City and Town because of lower proportion of mothers aged 35 and over.

### Population prevalence of Down’s syndrome in China in 2012

The total number of live births was available from 1985 to 2012 (in terms of millions, Table 1), but the maternal age distribution was not available for some years because no surveys were conducted in those years. Therefore we replaced the DS prevalence of the year when that year was not available with the data from the previous year. Table 1 shows the estimated number of people born with Down’s syndrome each year and the probability of those people survived to 2012. The total number of people with Down’s syndrome survived to 2012 was calculated in the last column in Table 1.

**Table 1:**
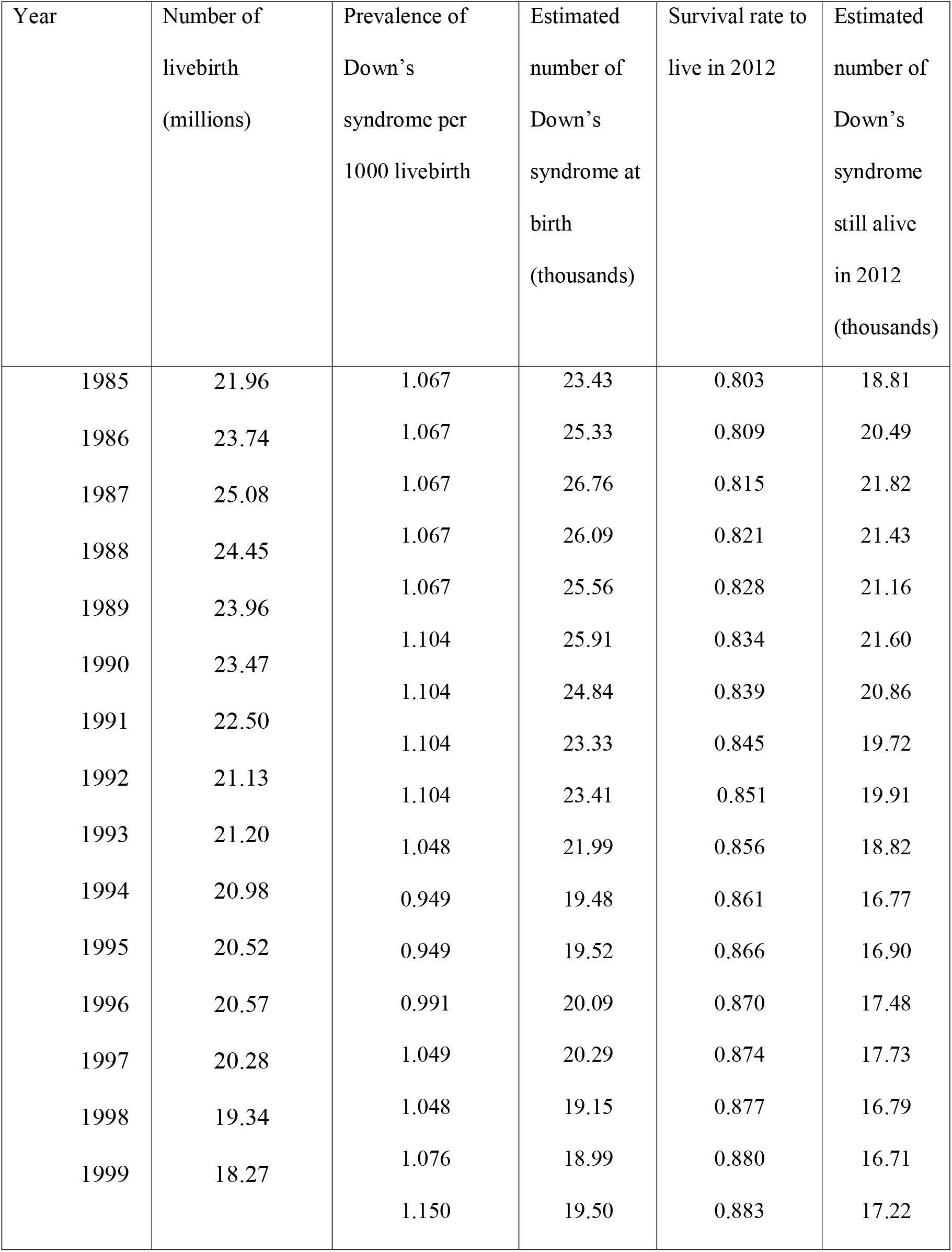
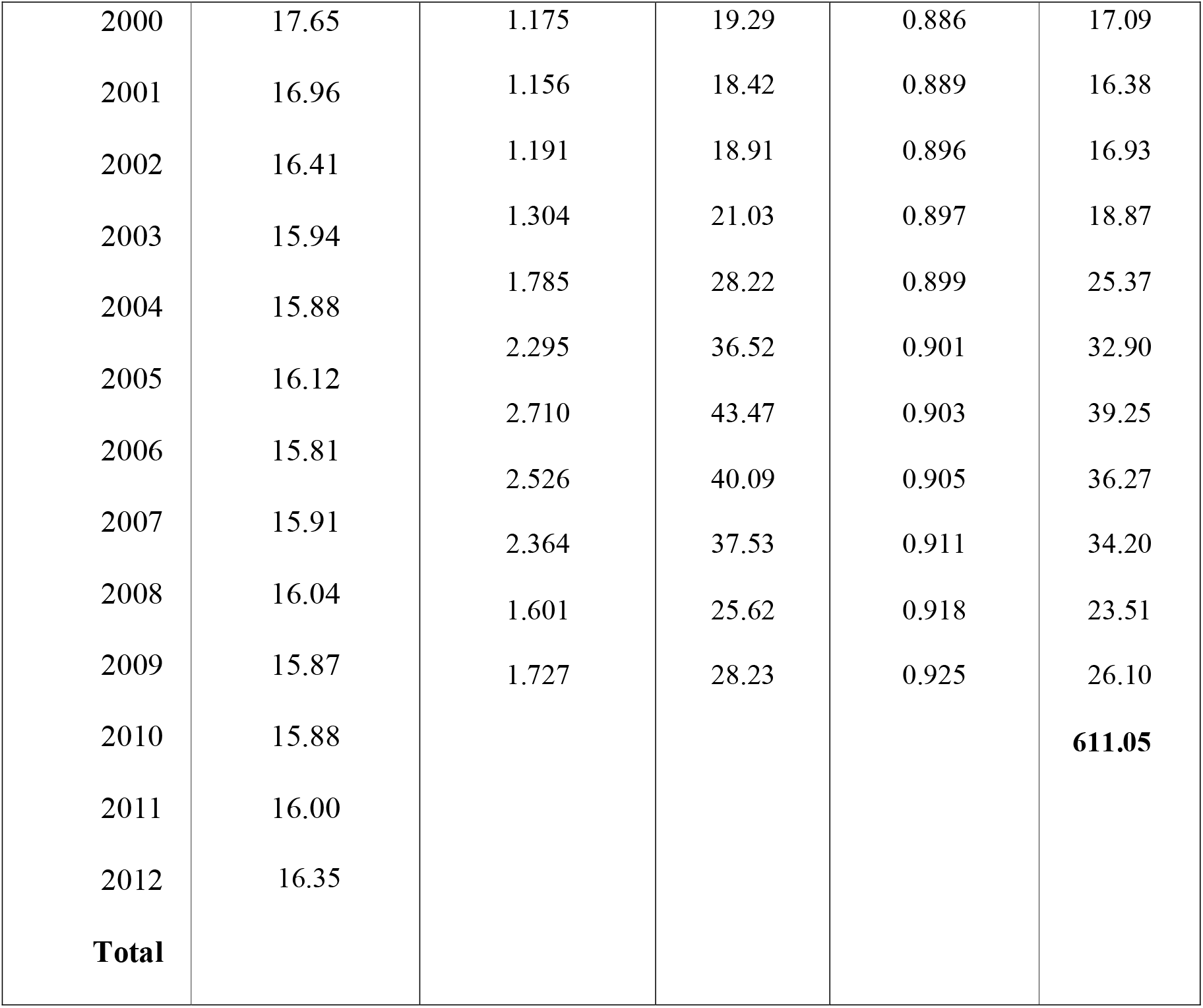
Estimated number of livebirths of Down’s syndrome from 1985 to 2012 in China and number of people with Down’s syndrome who was still alive in 2012

In total, there were estimated to be 611,053 people with Down’s syndrome aged 27 and below in China in 2012. Due to unavailability of national maternal age distribution prior to 1985, we cannot estimate the number of people with Down syndrome who were born before 1985. The estimated population prevalence of Down’s syndrome was 0.451 per 1000 (calculated by 611,053 divided by 1.35 billion of total population) (8).

In total, there were estimated to be 611,053 people with Down’s syndrome aged 27 and below in China in 2012. Due to unavailability of national maternal age distribution prior to 1985, we cannot estimate the number of people with Down syndrome who were born before 1985. The estimated population prevalence of Down’s syndrome was 0.451 per 1000 (calculated by 611,053 divided by 1,354,040,000 of total national population).(8)

## DISCUSSIONS

This paper presented the trends of maternal age distribution at national and regional levels between 1985 and 2012, and estimated the live birth prevalence of Down’s syndrome in China in that period. The live birth data used in this paper was from the annual population survey conducted by the National Bureau of Statistics since 1985, and deemed as representative birth data of the year.

Maternal age is the main predictor for Down’s syndrome, and the distribution of maternal age has changed significantly over the past 30 years at national and regional levels in China. The proportion of mothers aged over 35 has increased from 3.62% in 1985 to 14.40% in 2010 nationally, resulting a significant increase of the live birth prevalence of Down’s syndrome in recent years. The estimated live birth prevalence of 1.2 per 1000 live births before 2005 was in line with the actual live birth prevalence in some cities and towns in the same time period reported in other studies (9-13). Moreover, our study shows that the birth prevalence of Down’s syndrome has kept increasing significantly since 2005 because of the changing pattern of the maternal age in recent years. The change of distribution in maternal age is more significant in City than in Town or County. That is probably because higher proportion of women has delayed to have children in City because of career and financial pressure, and recent change in allowing second-child policy would further increase the proportion of mothers aged 35 and over, especially in City (14).

The surge of the estimated birth prevalence of Down’s syndrome was partly due to the increasing proportion of mothers aged 35 and over, but the other significant contributing factor was the heavy tail of the maternal distribution. We plotted the proportion of live birth against maternal age for 2000 and 2008 in China in red curves in Figure 4. The proportion of live birth decreased much slower with age in 2008 than in 2000. Meanwhile, we compared the distribution of maternal age in China with the data from England in the same time period. The shape of the maternal age distribution in England was more symmetric and bell shaped with peak around the maternal age of 29. However the shape of maternal age distribution for Chinese is right-skewed with heavier tail and peaks at 24. The differences in maternal age distribution between countries and across time periods had significant effect on the estimate of the live birth prevalence of Down’s syndrome. For example, the birth prevalence without prenatal screening and subsequent termination in England was 2.1 per 1000 live births comparing to 2.7 per 1000 live births in China in 2008, but the proportion of mothers aged 35 and over was 18% in England comparing to 16.7% in China (1, 2). The difference was largely due to the heavier tail in the maternal age distribution where maternal age was greater than 41 (Figure 4).

**Figure 4.**
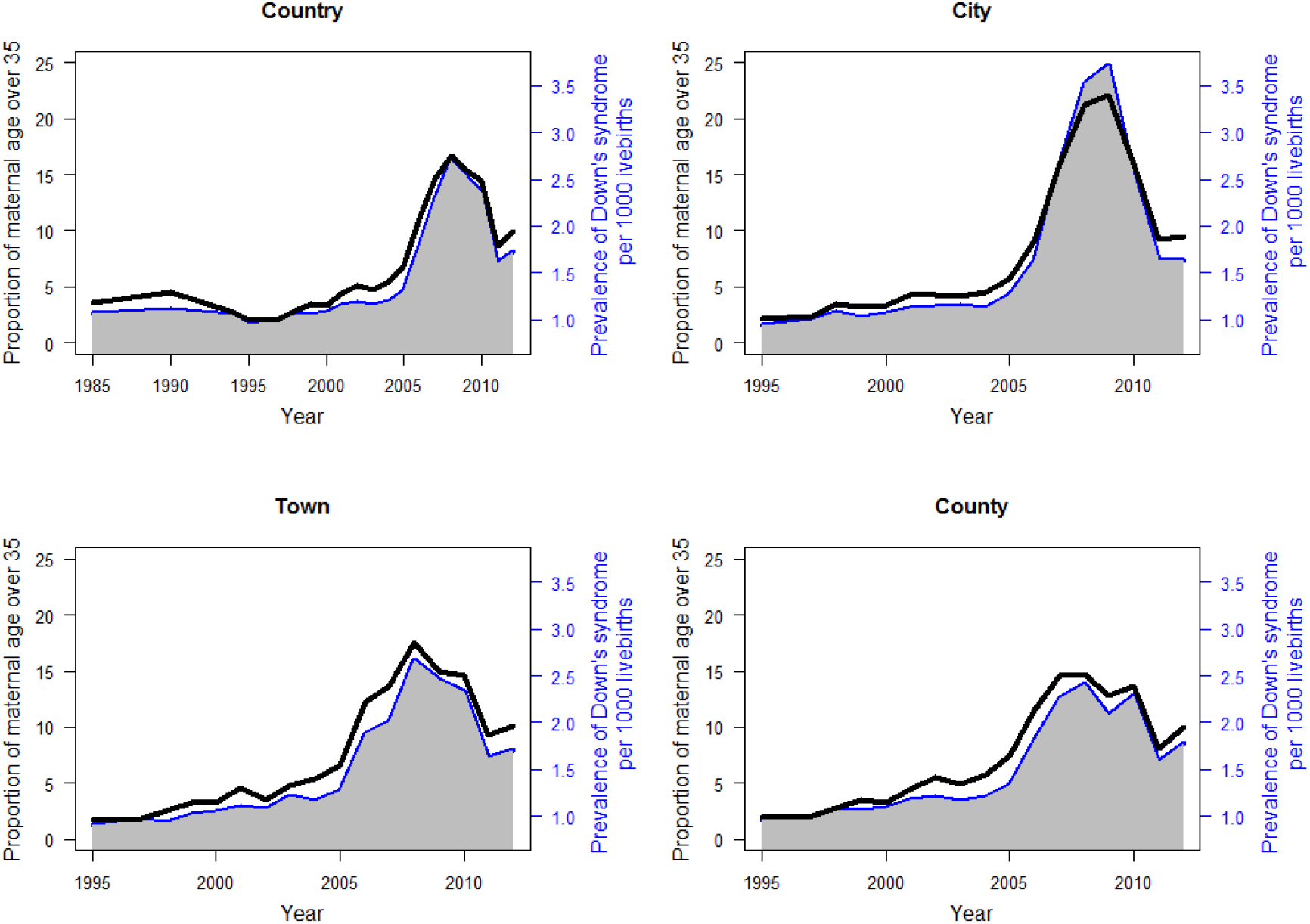
Comparison of maternal age distribution between China and England in 2000 and 2008.

The live birth prevalence of Down’s syndrome estimated in this paper could be different from the actual birth prevalence because of prenatal screening and subsequent termination implemented since 1990s. Many urban areas in China, especially those economically developed cities such as Beijing or Shanghai have included the Karyotypic confirmation of Down’s syndrome in the prenatal care for pregnant women in recent years(9, 14, 15). However, in rural areas where such medical devices and cares are not as accessible as in urban areas, the early detection of Down’s syndrome in perinatal birth is difficult not to mention the prenatal screening and termination(11). The estimated birth prevalence of Down’s syndrome presented in this paper has already given a clear rising trend of prevalence by urban and rural areas, but the actual difference in birth prevalence was difficult to estimate due to lack of data with prenatal screening and termination.

There are a few limitations of the study. Firstly, the association risk equation (1) between the risk of Down’s syndrome and maternal age was developed in the western countries (mainly Caucasian population). It may not be accurate to apply to Chinese population, although it has been investigated in other studies that the live birth prevalence in Chinese population is not significantly different from that in Caucasian population(16, 17). Secondly, the estimated birth prevalence of Down’s syndrome does not take into account of the prenatal screening and subsequent termination. The estimated prevalence would be significantly higher than actual prevalence if the termination rate is high (14, 18). Thirdly, the actual survival rates for people with Down’s syndrome in Chinese population could be much worse than those used in the paper, as no survival rate for DS people in China has been reported and the survival rate was adapted from western population which have better access to medical facilities and care.

In conclusion, this paper provides a clear message about the rising trend in maternal age distribution in both urban and rural areas in China and its effect on the estimated birth prevalence of babies with Down’s syndrome. The regional differences should be taken into account for planning population policy and allocating medical resources to reduce the burden of family with Down’s syndrome.

## Data Availability

The data used in this study is from National Bureau of Statistics of the People's Republic of China: http://www.stats.gov.cn/tjsj/ndsj/#
and from China statistics yearbook: http://nianjian.xiaze.com/plus/download.php?open=2&id=18067&uhash=b1b488b63840640a0b40304c 

http://www.stats.gov.cn/tjsj/ndsj/#

## Supplement

**Supplement Figure 1:**
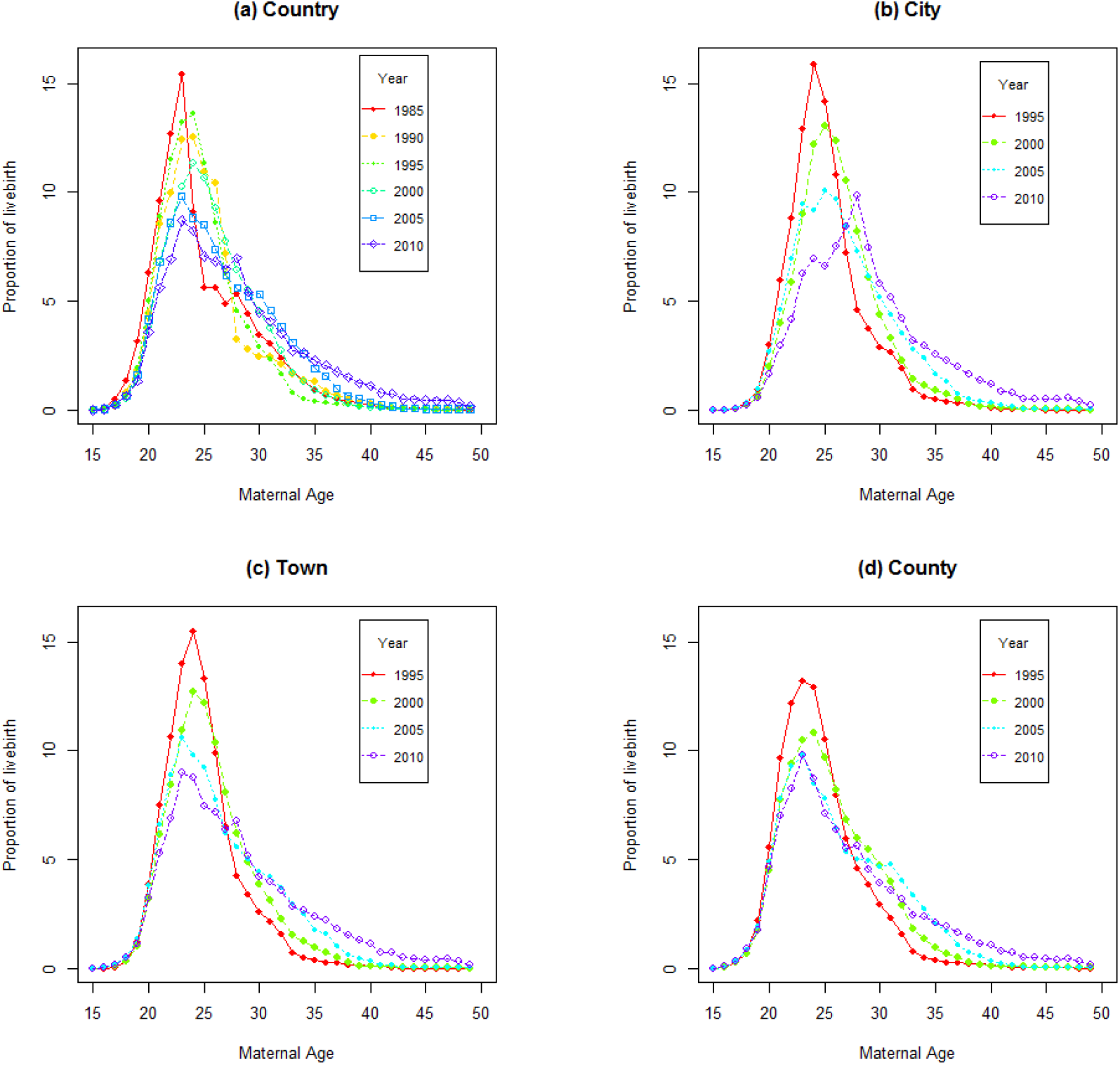
Maternal age distribution in China between 1985 and 2010 at national scale (a) and in different administrative regions (city, town and county, panels b-d). shows the detailed maternal age distribution in China between 198 and 2010 at national and regional levels. The maternal age has shifted towards older age from 1985 (red curve in figure 1a) to 2010 (purple curve in figure 1a). Such shift was more significant for mothers in City than in Town and County (figure 1b-d).

**Supplement Table 1:**
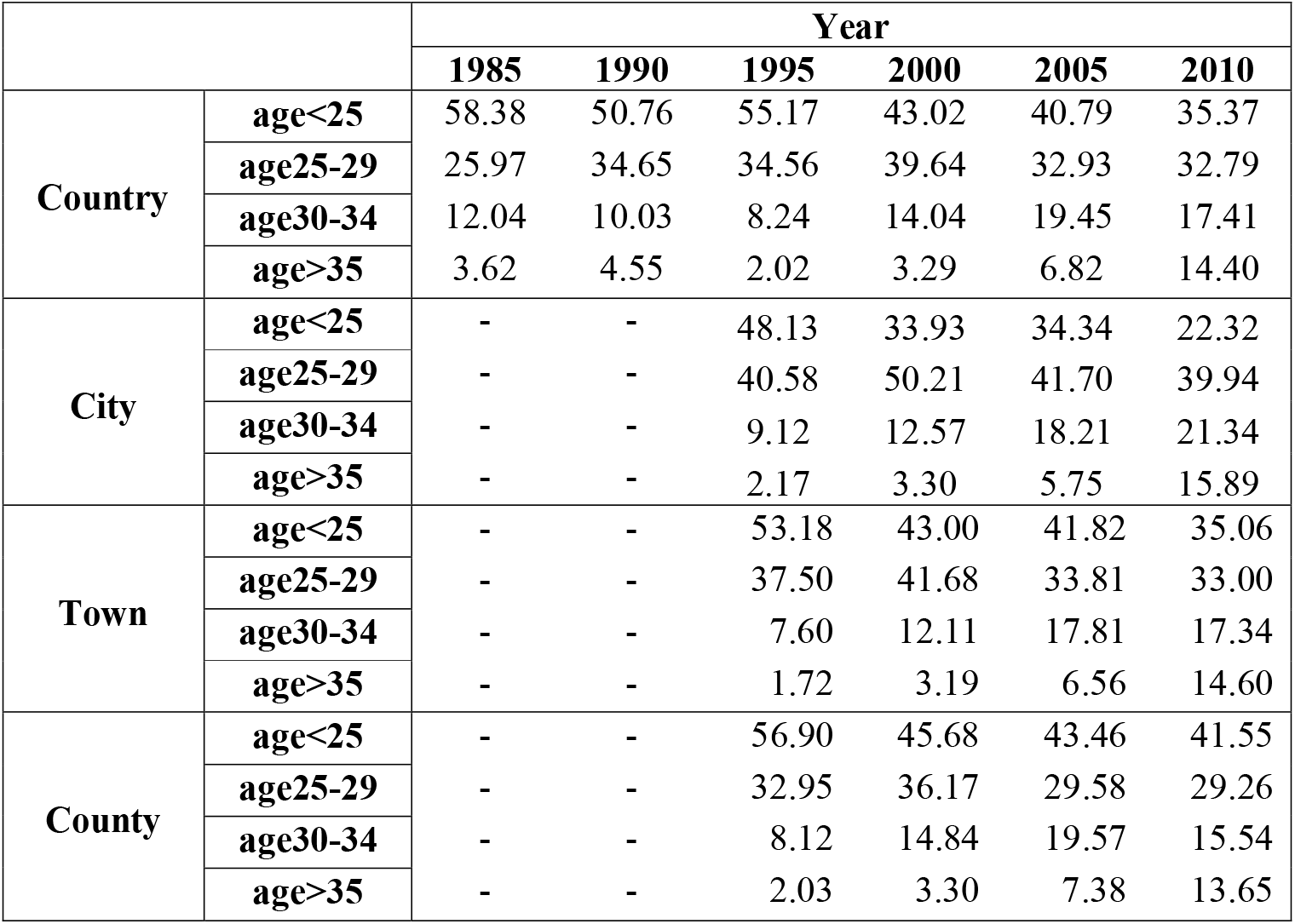
Proportion (per 100) of maternal age groups for country and regional levels for selected years, corresponding to Figure 2 in main text.

